# Investigating SARS-CoV-2 surface and air contamination in an acute healthcare setting during the peak of the COVID-19 pandemic in London

**DOI:** 10.1101/2020.05.24.20110346

**Authors:** Jie Zhou, Jonathan A. Otter, James R. Price, Cristina Cimpeanu, Danel Meno Garcia, James Kinross, Piers R Boshier, Sam Mason, Frances Bolt, Alison H. Holmes, Wendy S. Barclay

## Abstract

**Background:** Evaluation of SARS-CoV-2 surface and air contamination during the COVID-19 pandemic in London.

**Methods:** We performed this prospective cross-sectional observational study in a multi-site London hospital. Air and surface samples were collected from seven clinical areas, occupied by patients with COVID-19, and a public area of the hospital. Three or four 1.0 m^3^ air samples were collected in each area using an active air sampler. Surface samples were collected by swabbing items in the immediate vicinity of each air sample. SARS-CoV-2 was detected by RT-qPCR and viral culture; the limit of detection for culturing SARS-CoV-2 from surfaces was determined.

**Results:** Viral RNA was detected on 114/218 (52.3%) of surfaces and 14/31 (38.7%) air samples but no virus was cultured. The proportion of surface samples contaminated with viral RNA varied by item sampled and by clinical area. Viral RNA was detected on surfaces i and in air in public areas of the hospital but was more likely to be found in areas immediately occupied by COVID-19 patients than in other areas (67/105 (63.8%) vs. 29/64 (45.3%) (odds ratio 0.5, 95% confidence interval 0.2-0.9, p=0.025, Chi squared test)). The high PCR Ct value for all samples (>30) indicated that the virus would not be culturable.

**Conclusions:** Our findings of extensive viral RNA contamination of surfaces and air across a range of acute healthcare settings in the absence of cultured virus underlines the potential risk from environmental contamination in managing COVID-19, and the need for effective use of PPE, physical distancing, and hand/surface hygiene.

## INTRODUCTION

Since it was identified in Wuhan, China, in late 2019, the severe acute respiratory syndrome coronavirus (SARS-CoV-2) has rapidly spread around the world, resulting in a coronavirus disease 2019 (COVID-19) pandemic.[1] Experience from previous SARS and influenza outbreaks and emerging evidence for SARS-CoV-2 suggests droplet and contact spread as primary transmission routes.[1, 2] Additionally, there is evidence that airborne spread can occur during aerosol generating procedures.[1, 2]

In-hospital transmission to patients and healthcare workers was a key feature of SARS-CoV-1. [1, 3] Hospital-onset COVID-19 infection has been reported, probably due to inadequate implementation of effective infection prevention and control measures.[4] The dynamics of transmission in the health care environment are unclear and likely to be multifactorial. Contaminated surfaces and air are recognised as a key part of the transmission dynamic of SARS, MERS, influenza, and other organisms in hospitals.[1, 2, 5] Laboratory evidence suggests that the SARS-CoV-2 virus can survive on dry surfaces and in aerosols for days to weeks, particularly on non-porous surfaces.[6, 7] Furthermore, SARS-CoV-2 RNA has been detected on surfaces and in the air in hospitals that are caring for patients with COVID-19.[8-16]

However, our understanding of the role of surface and air contamination in the transmission of SARS-CoV-2 is limited. Most studies to date have relied on PCR to detect SARS-CoV-2 on surfaces and in air, and have not attempted to culture live virus thereby limiting the ability to interpret the relevance of detection by PCR; most studies published so far have focussed upon one geographical region (Asia), and included a limited selection of clinical and non-clinical areas were included with no evidence from operating theatre environments.[8, 9, 11, 12, 14, 15] In mid-April 2020, the UK was experiencing the first wave of the COVID-19 pandemic. During this period, there was evidence for hospital acquired infections with COVID-19.[17] Therefore, to inform and optimise infection prevention and control interventions, we evaluated surface and air contamination across a range of clinically-relevant locations (including operating theatres) and public areas during the peak of the | COVID-19 pandemic in London, using both RT-PCR and viral culture to detect SARS-CoV-2. We also performed supporting laboratory experiments to provide evidence on the viability of SARS-CoV-2 on surfaces, with associated limits of detection to qualify our findings.

## METHODS

### Setting

Sample collection for this prospective cross-sectional study was performed between April 2^nd^ and 20^th^ 2020 on selected wards at a large North West London teaching hospital group comprising five hospitals across four sites with 1,200 acute beds, which prior to the pandemic undertook 1.2 million episodes of patient contact per year. Most sampling was conducted on one hospital site during the peak of the COVID-19 pandemic (Supplemental Figure 1) when most patients were managed in cohort wards.

### Clinical areas selected for air and surface sampling

Seven clinical areas and a public area of the hospital were selected to represent a range of clinical environments within our hospital group. These included:

- Adult emergency department, which included sections dedicated for suspected and confirmed COVID-19 patients (with 19 cubicles and a 6-bedded resuscitation bay) and for patients not suspected to have COVID-19 (with a two cubicle-bay, and two four-cubicle bays).
- A 16-bedded COVID-19 cohorting adult acute admissions unit with four four-bedded bay.
- A 32-bedded COVID-19 cohorting adult intensive care unit with four four-bedded bays and 16 single rooms.
- Theatres during tracheostomy procedures.
- Two adult COVID-19 cohort wards: one 20-bed ward with four four-bedded bays and four single rooms, and one 19-bed ward with a nine-bedded bay, an 8 bedded-bay and two single rooms.
- An adult ward area including a 6-bedded bay converted into a negative pressure area for management of continuous positive airway pressure (CPAP) on patients with COVID-19.
- The entrance and public area of the main hospital building.

All inpatient wards were fully occupied by patients with COVID-19 at the time of sampling, apart from the Emergency Department. In the part of the Emergency Department dedicated for patients with confirmed or suspected COVID-19, two of the cubicles were occupied and one patient was in the ambulatory wait area at the time of sampling. These areas were disinfected daily using a combined chlorine-based detergent/disinfectant (Actichlor Plus, Ecolab), with an additional twice daily disinfection of high touch surfaces using the same detergent/disinfectant.

In each of these clinical areas, four air samples were collected (five air samples were collected in the Emergency Department, and three in public areas of the hospital). Surfaces in the immediate vicinity of each air sample that were considered to be touched frequently by staff or patients were sampled. These included bed rails, clinical monitoring devices (blood pressure monitors), ward telephones, computer keyboards, clinical equipment (syringe pumps, urinary catheters), hand-cleaning facilities (hand washing basins, alcohol gel dispensers). In each clinical area, sampling was performed in both patient (i.e. bays and single rooms) and non-patient care areas (i.e.nursing stations and staff rooms).

Environmental sampling was conducted during three tracheostomy procedures. During the first procedure, air sampling was performed before and during the procedure; for the other procedures, air sampling was performed during the procedure only.

### Sampling methods

Air sampling was performed using a Coriolis μ air sampler (referred to as Coriolis hereafter) (Bertin Technologies), which collects air at 100-300 - litres per minute (LPM). After 10 min sampling at 100 LPM, a total of 1.0 m^3^ air was sampled into a conical vial containing 5 mL; Dulbeccos’s minimal essential medium (DMEM). Surface samples were collected by swabbing approximately 25 cm^2^ areas of each item using flocked swabs (Copan, US) moistened in DMEM. Temperature, humidity and time of day were recorded at the time of sampling. In all clinical settings, samples were taken in order from the lowest to highest perceived risk of SARS-CoV-2 contamination.

### Detection and quantification of SARS-CoV-2 viral RNA genome and viral culture

Viral RNA detection and absolute quantification was performed using quantitative real-time reverse transcription polymerase chain reaction (RT-qPCR). Samples were extracted from 140 μL of the DMEM medium using the QIAamp viral RNA mini Kit according to the manufacturer’s instructions (Qiagen, Germany). Negative controls (water) were extracted and included in the PCR assays. SARS-CoV-2 viral RNA was detected using AgPath-ID One-Step RT-PCR Reagents (Life Technologies) with specific primers and probes targeting the envelop (E) gene.[18] The number of SARS-CoV-2 virus E gene copies per m^3^ air and copies per swab were calculated. All samples were run in duplicate.

Viral culture: Vero E6 (African Green monkey kidney) and Caco2 (human colon carcinoma) cells were used to culture virus from air and environmental samples. The cells were cultured in DMEM supplemented with heat inactivated fetal bovine serum (10%) and Penicillin/Streptomycin (10, 000 IU/mL &10, 000 μg/mL). For propagation, 200 μL of samples were added to 24 well plates. After 5-7 days, cell supernatants were collected, and RT-qPCR to detect SARS-CoV-2 performed as described above. Samples with at least one log increase in copy numbers for the E gene (reduced Ct values relative to the original samples) after propagation in cells were considered positive by viral culture.

We defined samples where both of the PCRs performed from an air or surface sample detected SARS-CoV-2 RNA as positive, and samples where one of the two PCRs performed from an air or surface sample detected SARS-CoV-2 RNA as suspected.

We performed a laboratory experiment to determine the limit of detection for culturing SARS-CoV-2 dried on surfaces. A dilution series from solution containing 8.25×10^6^ PFU/mL SARS-CoV-2 (titred by plaque assay in Vero cells) from 10^-3^ to 10^-6^ (covering Ct values from 26 to 36) was produced in DMEM and 50 μL inoculated in triplicate onto the surface of plastic (standard keyboard key) or stainless steel (2 × 1 × 0.2 cm) pieces. The inoculated surfaces were dried in a safety cabinet for 2 hours after which they were visibly dry. They were then sampled using flocked swabs. Swabs were deposited into 1.5 mL of DMEM for 1 hour. RT-qPCR was used to determine viability following 7 days of culture as follows. 140 μL was used for RNA extraction and qPCR immediately (0 days post inoculation, dpi) and after incubation for 7 days in a 24-well plate with VeroE6 cells (7 dpi). Samples with an increase in copy numbers for the E gene (reduced Ct values relative to the original samples) after propagation in Vero E6 cells were considered positive by viral culture.

## Statistical analysis

A Chi square test was used to compare the proportion of environmental samples (surfaces or air) that were positive or suspected for SARS-CoV-2 RNA in areas immediately occupied by patients with COVID-19 with other areas. The mean concentration of air and surface contamination in each of the areas was log transformed and then compared by one-way ANOVA followed by Tukey’s multiple comparisons test.

### Ethics approval

The work was registered locally as an NHS service evaluation (#434).

## RESULTS

114/218 (52.3%) of surface samples were suspected (91/218 (41.7%)) or positive (23/218 (10.6%)) for SARS-CoV-2 RNA but no virus was cultured (Table 1). The proportion of surface samples contaminated varied by item, with suspected or positive RNA samples identified on >80% of computer keyboards/mice, alcohol gel dispensers, and chairs, and >50% of toilet seats, sink taps, and patient bedrails (Figure 1). A similar pattern was observed in air samples; no virus was cultured, but 14/31 (38.7%) of samples were suspected (12/31 (38.7%)) or positive 92/31 (6.4%)) for SARS-CoV-2 RNA (Table 1).

**Table 1.** PCR results from surface and air samples.

|  |  | Total | SURFACE SAMPLES |  |  |  |  |  | Result | AIR SAMPLES |  |
| --- | --- | --- | --- | --- | --- | --- | --- | --- | --- | --- | --- |
|  |  |  | positive | %positive | suspect | %suspect | positive or suspect | % positive or suspect |  | Concentration (copies/m<sup>3</sup>) | Notes |
| **Cohort ward A** | Staff room | 6 | 0 | 0.0 | 2 | 33.3 | 2 | 33.3 | Negative |  |  |
|  | Nurse station | 6 | 1 | 16.7 | 3 | 50.0 | 4 | 66.7 | Negative |  |  |
|  | Toilet B (outside the patients' bay) | 6 | 0 | 0.0 | 2 | 33.3 | 2 | 33.3 | Negative |  |  |
|  | Cohort bay B | 6 | 3 | 50.0 | 2 | 33.3 | 5 | 83.3 | Positive | 7048 |  |
| **Cohort ward B** | Staff room | 4 | 0 | 0.0 | 0 | 0.0 | 0 | 0.0 | Negative |  |  |
|  | Patients' toilet (in the ward) | 7 | 0 | 0.0 | 1 | 14.3 | 1 | 14.3 | Suspect | 464 |  |
|  | Male bay | 12 | 1 | 8.3 | 4 | 33.3 | 5 | 41.7 | Suspect | 1335 |  |
|  | Male bay (side room) | 8 | 2 | 25.0 | 5 | 62.5 | 7 | 87.5 | Suspect | 163 |  |
| **Adult acute admission unit** | Ward managers office | 5 | 1 | 20.0 | 2 | 40.0 | 3 | 60.0 | Negative |  |  |
|  | Nurse station | 7 | 0 | 0.0 | 5 | 71.4 | 5 | 71.4 | Positive | 404 |  |
|  | Patient bay 2 | 8 | 0 | 0.0 | 2 | 25.0 | 2 | 25.0 | Negative |  |  |
|  | Patient bay 1 | 10 | 0 | 0.0 | 8 | 80.0 | 8 | 80.0 | Negative |  |  |
| **Adult emergency department** | 'Green' majors | 10 | 1 | 10.0 | 5 | 50.0 | 6 | 60.0 | Negative |  |  |
|  | Nurse station | 4 | 2 | 50.0 | 0 | 0.0 | 2 | 50.0 | Negative |  |  |
|  | Ambulatory waiting | 3 | 2 | 66.7 | 1 | 33.3 | 3 | 100.0 | Negative |  |  |
|  | Patient assessment cubicles | 3 | 0 | 0.0 | 1 | 33.3 | 1 | 33.3 |  |  |  |
|  | Male toilet (next to the nurse station) | 2 | 0 | 0.0 | 1 | 50.0 | 1 | 50.0 |  |  |  |
|  | Resus bay (last patient > 2 hours) | 10 | 0 | 0.0 | 4 | 40.0 | 4 | 40.0 | Suspect | 35 |  |
| **Hospital public areas** | QEQM main entrance | 7 | 1 | 14.3 | 4 | 57.1 | 5 | 71.4 | Suspect | 1574 |  |
|  | Male toilet at QEQM main entrance | 7 | 1 | 14.3 | 3 | 42.9 | 4 | 57.1 | Suspect | 1545 |  |
|  | Lift area QEQM ground floor | 10 | 0 | 0.0 | 4 | 40.0 | 4 | 40.0 | Negative |  |  |
| **Temporary CPAP ward** | Nurse station | 5 | 1 | 20.0 | 2 | 40.0 | 3 | 60.0 | Suspect | 1922 |  |
|  | CPAP unit | 19 | 2 | 10.5 | 12 | 63.2 | 14 | 73.7 | Suspect | 31 | < 1m from 2 patients |
|  |  |  |  |  |  |  |  |  | Negative |  | > 2 m from patients |
|  | PPE doffing area | 5 | 0 | 0.0 | 2 | 40.0 | 2 | 40.0 | Negative |  |  |
| **Adult ICU** | Staff room | 10 | 0 | 0.0 | 6 | 60.0 | 6 | 60.0 | Suspect | 249 |  |
|  | Nurse station inside ICU | 6 | 1 | 16.7 | 0 | 0.0 | 1 | 16.7 | Negative |  |  |
|  | Bay area | 11 | 0 | 0.0 | 5 | 45.5 | 5 | 45.5 | Suspect | 164 |  |
|  | Side room bay area | 8 | 2 | 25.0 | 4 | 50.0 | 6 | 75.0 | Suspect | 307 |  |
| **Theatres** | Theatres | 13 | 2 | 15.4 | 1 | 7.7 | 3 | 23.1 | Negative |  | Before tracheostomy |
|  |  |  |  |  |  |  |  |  | Negative |  | During tracheostomy |
|  |  |  |  |  |  |  |  |  | Suspect | 1163 | During tracheostomy |
|  |  |  |  |  |  |  |  |  | Negative |  | During tracheostomy |
| **Total** |  | **218** | **23** | **10.6** | **91** | **41.7** | **114** | **52.3** | **2/31 (6.4%) positive; 12/31 (38.7%) suspect** |  |  |

**Figure 1.**
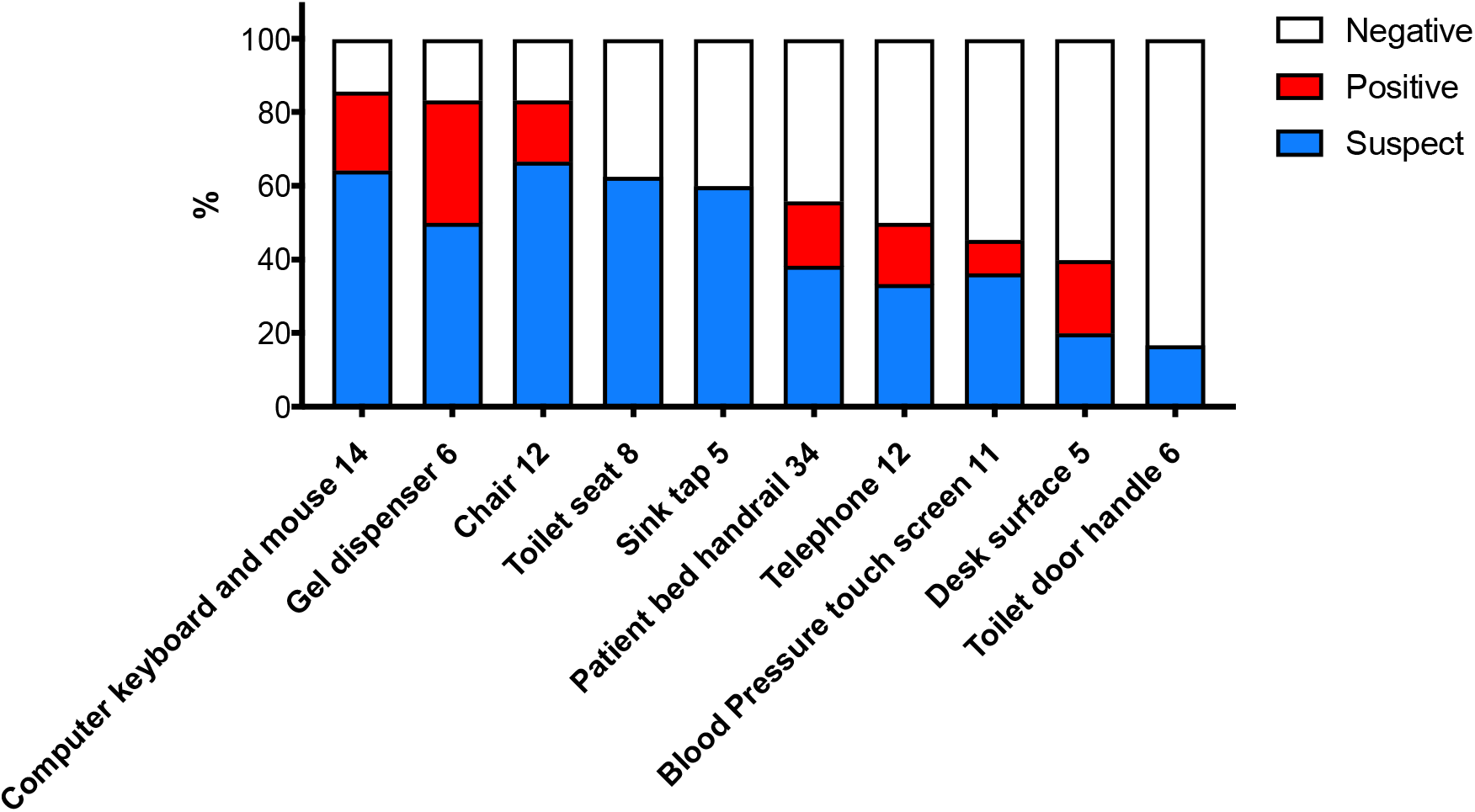
Proportion of environmental samples suspected or positive by item sampled. The number of the x axis represented the number of each item sampled.

SARS-CoV-2 RNA was detected in air samples from all eight areas tested with levels ranging from 10^1^ to 10^3^ genome copies / m^3^ (Table 1); there was no significant difference in mean viral RNA concentration across the eight areas tested (p=0.826). Similarly, SARS-CoV-2 RNA was detected in surface samples from all eight areas tested, with levels ranging from 10^1^ to 10^4^ copies per swab (Figure 2). There was a significant difference in the mean SARS-CoV-2 surface viral load across the eight areas tested (p=0.004), with both Cohort Ward A and the Temporary CPAP ward showing higher levels of viral RNA; Cohort Ward A (mean = 1.76 log_10_ copies/swab) > Adult ICU (mean = 0.0018 log_10_ copies/swab) (p = 0.015), and the Temporary CPAP Ward (mean = 1.69 log_10_ copies/swab) > Adult ICU (p = 0.016).

**Figure 2.**
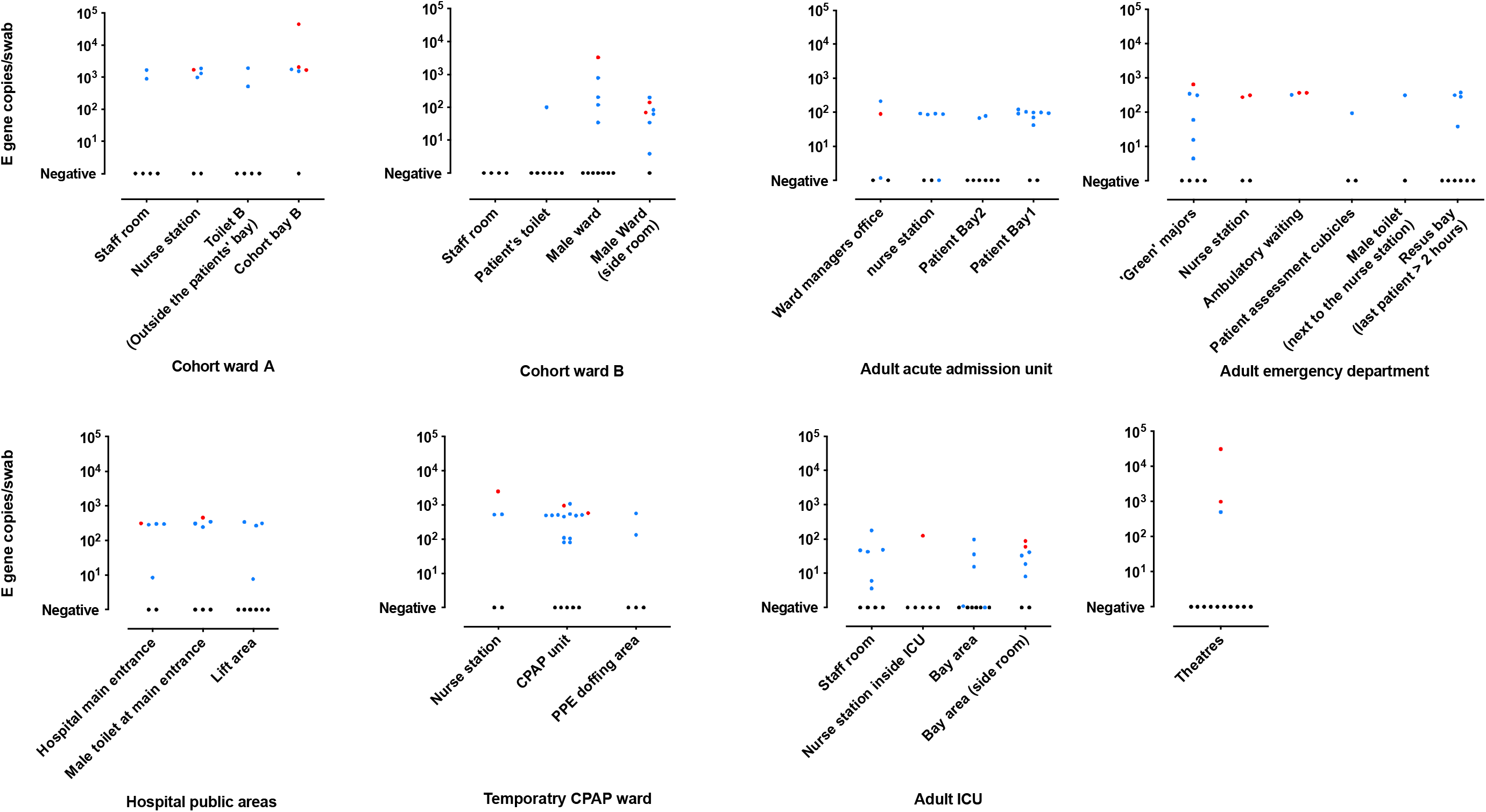
SARS-CoV-19 E gene copy number from surface swabs. The quantity of E gene copy number per swab is shown. Suspect samples = blue dots; positive samples = red dots; negative samples = black dots.

Several clinical areas where AGPs are commonly performed were sampled. A suspected positive air sample was collected in the resuscitation bay in the emergency department, where aerosol generating procedures are commonly performed (although had not been performed for more than two hours prior to sample collection). In a ward temporarily converted for CPAP, SARS-CoV-2 RNA was detected from air within the negative pressure CPAP bay, and outside the bay. No patient was undergoing CPAP at the time of sampling, but one patient was undergoing high-flow nasal cannula (HFNC) oxygen therapy. In the adult ICU, 3/4 air samples were suspected or positive. In operating theatres, 1/3 air samples collecting during three tracheostomy procedures was positive.

SARS-CoV-2 RNA was detected in surface and air samples in parts of the hospital hosting staff but not being used for direct patient care, including the staff room in the ICU, the nursing station outside of the CPAP unit, and in the hospital main entrance and public toilets. However, positive or suspected air and surface samples were significantly more likely to be found in areas immediately occupied by COVID-19 patients than in other areas (67/105 (63.8%) in areas immediately occupied by COVID-19 patients vs. 29/64 (45.3%) in other areas (odds ratio 0.5, 95% confidence interval 0.2-0.9, p=0.025).

Since viable virus was not cultured from any of the air or surface samples, we performed laboratory experiments to determine the limit of detection of SARS-CoV-2 dried onto i surfaces. Viable SARS-CoV-2 virus could be cultured from experimentally contaminated dried surfaces with a Ct value <30; this was consistent for plastic and metal test surfaces (Table 2). In our study, all surface and air samples from the hospital environment had a Ct value >30.

**Table 2:** Viability of SARS-CoV-2 dried onto steel or plastic surfaces from a dilution series; viability determined through RT-PCR from cultures immediately after drying, 0 days post inoculation (dpi) with Vero E6 cells compared with after culture (7 dpi). Means and standard deviations of Ct values are shown.

| Inoculum (PFU) | Steel surface |  | Plastic surface |  |
| --- | --- | --- | --- | --- |
|  | After drying (Ct) | After culture (Ct) | After drying (Ct) | After culture (Ct) |
| 41.25 | 26.23 ± 0.30 | 12.65 ± 0.51 Pos | 25.95 ± 0.06 | 11.16 ± 0.19 Pos |
| 4.125 | 29.27 ± 0.04 | 12.86 ± 0.01 Pos | 29.51 ± 0.29 | 12.58 ± 1.47 Pos |
| 0.4125 | 32.54 ± 0.06 | 36.48 ± 1.80 Neg | 32.67 ± 0.07 | 37.39 ± 0.21 Neg |
| 0.04125 | 39.22 ± 5.13 | 41.33 ± 3.45 Neg | 36.55 ± 0.23 | 39.76 ± 4.61 Neg |

## DISCUSSION

SARS-CoV-2 RNA was detected frequently from surface and air samples but we did not identify viable virus in any surface or air sample. Furthermore, our simulated laboratory studies showed that the RNA levels detected on environmental surfaces in the hospital were lower than the minimum that can be cultured from surfaces two hours after virus is deposited. SARS-CoV-2 RNA was identified across the eight areas that we tested, including areas of the hospital not used to care for patients with COVID-19 (e.g. public areas of the hospital). However surface and air contamination was significantly more frequent in areas immediately occupied by COVID-19 patients than in other areas.

A direct comparison between our findings and other studies that have evaluated contamination of surfaces and air with SARS-CoV-2 is not possible due to differences in: environmental sampling strategy (including which clinical areas were included, which surfaces were sampled, and where air samples were collected from); experimental methods (including the method for sampling surfaces and the sampler used for air); the phase of the pandemic during which sampling was performed; the physical layout of buildings and clinical spaces (including the efficiency of air handling systems); individual patient characteristics that have been shonwn to influence shedding of SARS-CoV-2 and other hospital pathogens including the stage and severity of disease and site of infection;[4, 19] and the patient and staff testing, and cleaning and disinfection protocols. Nonetheless, our finding of widespread detection of viral RNA on surfaces (114/218, 52.3%) and to a lesser extent air (14/31, 38.7%) is broadly consistent with the findings of most others although the proportion of surface and air samples positive for viral RNA is higher in our study.[8-13] For example, Ye et al. performed PCR detection of surface contamination in a range of clinical settings in a | hospital caring for patients with COVID-19 in Wuhan, China.[9] Overall, 14% of 626 surface samples were positive for viral RNA, with a higher proportion of surface samples positive in the ICU (32% of 69 samples). However, other studies have identified very little or no contamination of surfaces or air.[8, 10] Other studies have observed higher frequencies of contamination in patient-care vs. non-patient-care areas,[8, 9, 11] and variation in the frequency of contamination across different clinical areas, which is in line with our findings.[9, 11] One surprising finding in our study was that the level of contamination on surfaces in the ICU was lower than in a cohort general ward or in the temporary CPAP ward, in contrast to other findings.[9] This may be because patients sampled in the ICU were on closed circuit ventilation systems through cuffed endotracheal tubes, which may have a lower risk of producing surface and air contamination than other ventilation systems such as CPAP.

We did not identify viable virus on any surface or air sample. Few studies have attempted to culture SAR-CoV-2 from healthcare environments, and no viable virus was detected.[10, 14] Our laboratory study of the viability of virus dried on surfaces helps to qualify our findings and the findings of others, suggesting that Ct values of >30 are unlikely to be culturable. Bearing in mind that the viral RNA detected in the hospital setting might have been deposited more than two hours previously, we cannot differentiate whether our inability to culture virus from the samples is explained by the low RNA levels or the length of time since deposition or both. It is also possible that virus was infectious but not culturable in the laboratory.

Surface contamination was detected on a range of items. Computer keyboards, chairs, and alcohol dispensers had the highest proportion of positive/suspected SAS-CoV-2 samples. Other studies have also identified computer keyboards and/or mice as a risk for contamination with SARS-CoV-2 RNA.[8, 9, 11] Many of the computers that we sampled were in shared staff clinical areas (such as nursing stations), so this argues for frequent disinfection of these items. The contamination of alcohol gel dispensers is unsurprising since staff hands activate these before hand hygiene is performed. However, alcohol gel dispensers should be included in routine cleaning and disinfection protocols or designed such that they can be activated without touching.

We sampled several areas where aerosol generating procedures are commonly performed including the resuscitation bay in the emergency department, ICU, temporary CPAP ward, and operating theatres during tracheostomies. Positive or suspected air samples were identified in all of these clinical areas at a level of 10^1^ to 10^3^ copies / m^3^. There was no difference in the viral load of the air across the eight areas sampled, which provides some evidence that AGPs do not produce persistently high levels of air contamination. However, we did not sample the air over time, and our air sampling method did not differentiate particle size so we are unable to distinguish droplets from aerosols (< 5 μM). One recent study evaluated contamination of the air with SARS-CoV-2 in a permanent hospital and in a field hospital in Wuhan, China.[13] Viral culture was not performed, but viral RNA was identified a low levels (in the 10^1^-10^2^ range copies per m^3^) in patient care areas, and was not detected or detected in very low levels in public areas. Positive samples were identified in a range of particle sizes, including those <5 μM, which would typically be considered as aerosols.[2] It seems likely, therefore, that the positive and suspected air samples identified in our study included a range of particle sizes spanning 5 μM, particularly in areas where aerosol generating procedures are common.

Whilst we performed sampling in a temporary CPAP ward, no patient was undergoing CPAP at the time of sampling. However, one patient was undergoing HFNC during the time of sample, and air contamination was identified <1 m from this patient. A recent summary of evidence concludes that HFNC is a lower risk procedure in terms of aerosol generation than CPAP, which should be a topic for future studies.[20]

We identified contamination of surfaces and air during three tracheostomy procedures. Several studies and commentaries have evaluated the potential for various surgical procedures to produce aerosols for patients with COVID-19.[21-23] One study evaluated the spread of droplets during tracheostomies, although did not include sampling for SARS-CoV-2.[21] Whilst our methods did not include measurement of particle size, our findings highlight a potential theoretical risk of transmission of COVID-19 during these procedures. However, a larger sample size is required to understand this risk

Our study has important strengths and limitations. Strengths include our sampling strategy encompassing contemporaneous surface and air samples from a range of clinical services including both patient care and non-patient care areas, specifically, we included operating theatres and areas dedicated to known and potential AGPs; each sample was tested using PCR and also viral culture, and we performed laboratory viral culture experiments to quality our findings; the sampling was conducted during the peak of the pandemic (and so likely represents a worst-case scenario) in a European hospital group. Limitations include not collecting patient samples to better understand how our findings links to patient samples, particularly during tracheostomies and AGPs; no asymptomatic patient or staff testing ongoing at the time of sampling, which means patients and staff without known COVID-19 could have been shedding SARS-CoV-2 and this would explain the detection of SARS-CoV-2 RNA in non-patient care areas; challenges in interpreting the significance of samples with low viral loads,; a lack of resolution of particle sizes for contamination of the air; and no longitudinal sampling was performed so these findings represent a “snapshot”.

Our findings may have implications for future policy and guidelines. Most international guidelines recommend enhanced surfaces disinfection during the management of COVID-19. For example, Public Health England recommends enhanced disinfection using a chlorine-based disinfectant (or a disinfectant with effectiveness against coronaviruses).[24] Our finding of widespread RNA contamination of clinical areas used to care for patients with COVID-19 supports the need for enhanced disinfection. Physical distancing is recommended by most governments and personal protective equipment (PPE) is recommended during contact with patients with COVID-19 plus higher levels of PPE for performing aerosol generating procedures. Whilst we did not measure particle sizes during our air sampling, our findings highlight a potential role for contaminated air in the spread of COVID-19. Our finding of air contamination outside of clinical areas should be considered i when making respiratory PPE recommendations in healthcare settings.[25]

Whilst SAR-CoV-2 RNA was detected within healthcare environments, further research linking patient, staff and environmental samples is required to better understand transmission routes. Longitudinal environmental and clinical sampling across healthcare settings is required to understand risk factors associated with viral shedding and transmission. Our findings can be used to parameterise mathematical models of COVID-19 transmission. Finally, our methods can be used to assess the potential risk associated with various procedures including some surgical and other procedures such as CPAP and nebulisation of medications. Findings from these studies may prompt changes to PPE recommendations for specific procedures, and the implementation of various innovative tools and approaches to reduce viral shedding (such as “helmet CPAP”).[26-28]

Whilst SARS-CoV-2 RNA was detected in clinical and non-clinical areas, no viable virus was recovered. These results are in line with other studies which have identified viral RNA but no viable SARS-CoV-2 within healthcare environments. Our findings of extensive viral RNA contamination of surfaces and air across a range of acute healthcare settings in the absence of cultured virus underlines the potential risk from surface and air contamination in managing COVID-19, and the need for effective use of PPE, physical distancing, and hand/surface hygiene.

## Data Availability

All data related to this work will be shared upon request.

## ACKNOWLEDGEMENTS

We wish to acknowledge the staff teams and patients who supported this sampling during the peak of the challenges posed by this pandemic.

## Key points

The role of surface and air contamination in SARS-CoV-2 transmission was evaluated in a London hospital. Whilst SARS-CoV-2-RNA was detected no viable virus was recovered. This underlines the potential risk of environmental contamination and the need for effective IPC practices.

## Author contributions

All authors met the ICMJE criteria for authorship. JZ and JAO conceived the study, collected and analysed data, and wrote the manuscript; JRP conceived the study, collected data, and contributed to the manuscript; CP, DMG, PRB, SM collected data and contributed to the manuscript; FB, AHH, and ASB conceived the study, analysed data, and contributed to the manuscript. JAO is the study guarantor.

---

**Supplemental Figure 1:**
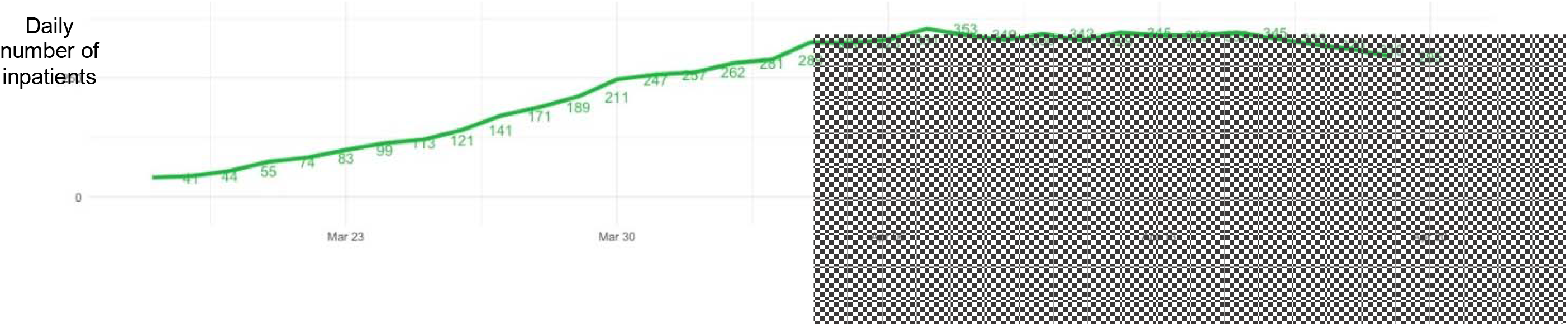
Trends in daily number of inpatients with COVID-19; the grey box indicates when surface and air samples were collected

## REFERENCES

1. Wilder-Smith A, Chiew CJ, Lee VJ. Can we contain the COVID-19 outbreak with the same measures as for SARS? The Lancet Infectious diseases, 2020; 20: e102-e7.

2. Otter JA, Donskey C, Yezli S, Douthwaite S, Goldenberg SD, Weber DJ. Transmission of SARS and MERS coronaviruses and influenza virus in healthcare settings: the possible role of dry surface contamination. The Journal of hospital infection, 2016; 92: 235–50.

3. Gowri G, Philip C, Yee Sin L, et al. SARS Transmission and Hospital Containment. Emerging Infectious Disease journal, 2004; 10: 395.

4. He X, Lau EHY, Wu P, et al. Temporal dynamics in viral shedding and transmissibility of COVID-19. Nature medicine, 2020; 26: 672–5.

5. Otter JA, Yezli S, Salkeld JA, French GL. Evidence that contaminated surfaces contribute to the transmission of hospital pathogens and an overview of strategies to address contaminated surfaces in hospital settings. Am J Infect Control, 2013; 41: S6-S11.

6. van Doremalen N, Bushmaker T, Morris DH, et al. Aerosol and Surface Stability of SARS-CoV-2 as Compared with SARS-CoV-1. The New England journal of medicine, 2020; 382: 1564–7.

7. Chin AWH, Chu JTS, Perera MRA, et al. Stability of SARS-CoV-2 in different environmental conditions. The Lancet Microbe, 2020; 1: e10.

8. Wu S, Wang Y, Jin X, Tian J, Liu J, Mao Y. Environmental contamination by SARS-CoV-2 in a designated hospital for coronavirus disease 2019. Am J Infect Control, 2020:

9. Ye G, Lin H, Chen S, et al. Environmental contamination of SARS-CoV-2 in healthcare premises. The Journal of infection, 2020:

10. Wang J, Feng H, Zhang S, et al. SARS-CoV-2 RNA detection of hospital isolation wards hygiene monitoring during the Coronavirus Disease 2019 outbreak in a Chinese hospital. International journal of infectious diseases: IJID: official publication of the International Society for Infectious Diseases, 2020; 94: 103–6.

11. Guo ZD, Wang ZY, Zhang SF, et al. Aerosol and Surface Distribution of Severe Acute Respiratory Syndrome Coronavirus 2 in Hospital Wards, Wuhan, China, 2020. Emerging infectious diseases, 2020; 26:

12. Ong SWX, Tan YK, Chia PY, et al. Air, Surface Environmental, and Personal Protective Equipment Contamination by Severe Acute Respiratory Syndrome Coronavirus 2 (SARS-CoV-2) From a Symptomatic Patient. Jama, 2020; 323: 1610–2.

13. Liu Y, Ning Z, Chen Y, et al. Aerodynamic analysis of SARS-CoV-2 in two Wuhan hospitals. Nature, 2020:

14. Colaneri M, Seminari E, Novati S, et al. SARS-CoV-2 RNA contamination of inanimate surfaces and virus viability in a health care emergency unit. Clinical microbiology and infection: the official publication of the European Society of Clinical Microbiology and Infectious Diseases, 2020:

15. Chia PY, Coleman KK, Tan YK, et al. Detection of air and surface contamination by SARS-CoV-2 in hospital rooms of infected patients. Nature communications, 2020; 11: 2800.

16. Santarpia JL, Rivera DN, Herrera V, et al. Transmission Potential of SARS-CoV-2 in Viral Shedding Observed at the University of Nebraska Medical Center. medRxiv, 2020: 2020.03.23.20039446.

17. Evans S, Agnew E, Vynnycky E, Robotham JV. The impact of testing and infection prevention and control strategies on within-hospital transmission dynamics of COVID-19 in English hospitals. *medRxiv*, 2020: 2020.05.12.20095562.

18. Corman VM, Landt O, Kaiser M, et al. Detection of 2019 novel coronavirus (2019-nCoV) by real-time RT-PCR. Euro surveillance: bulletin Europeen sur les maladies transmissibles = European communicable disease bulletin, 2020; 25:

19. Otter JA, Yezli S, French GL. The role played by contaminated surfaces in the transmission of nosocomial pathogens. Infection control and hospital epidemiology, 2011; 32: 687–99.

20. Li J, Fink JB, Ehrmann S. High-flow nasal cannula for COVID-19 patients: low risk of bio-aerosol dispersion. The European respiratory journal, 2020:

21. Chow VLY, Chan JYW, Ho VWY, et al. Tracheostomy during COVID-19 pandemic-Novel approach. Head & neck, 2020:

22. Thamboo A, Lea J, Sommer DD, et al. Clinical evidence based review and recommendations of aerosol generating medical procedures in otolaryngology - head and neck surgery during the COVID-19 pandemic. Journal of otolaryngology - head & neck surgery = Le Journal d’oto-rhino-laryngologie et de chirurgie cervico-faciale, 2020; 49: 28.

23. RN, Wong SH, Sánchez-Luna SA, et al. Overview of guidance for endoscopy during the coronavirus disease 2019 pandemic. Journal of gastroenterology and hepatology, 2020; 35: 749–59.

24. PHE. COVID-19: infection prevention and control guidance https://assets.publishing.service.gov.uk/government/uploads/svstem/uploads/attachmentdata/file/886668/COVID-19Infectionpreventionandcontrolguidancecomplete.pdf. 2020:

25. Garcia Godoy LR, Jones AE, Anderson TN, et al. Facial protection for healthcare workers during pandemics: a scoping review. BMJ global health, 2020; 5:

26. Radovanovic D, Rizzi M, Pini S, Saad M, Chiumello DA, Santus P. Helmet CPAP to Treat Acute Hypoxemic Respiratory Failure in Patients with COVID-19: A Management Strategy Proposal. Journal of clinical medicine, 2020; 9:

27. David AP, Jiam NT, Reither JM, Gurrola JG, 2nd, Aghi M, El-Sayed IH. Endoscopic Skull Base and Transoral Surgery During the COVID-19 Pandemic: Minimizing Droplet Spread with a Negative-Pressure Otolaryngology Viral Isolation Drape (NOVID). Head & neck, 2020:

28. Hirschmann MT, Hart A, Henckel J, Sadoghi P, Seil R, Mouton C. COVID-19 coronavirus: recommended personal protective equipment for the orthopaedic and trauma surgeon. Knee surgery, sports traumatology, arthroscopy: official journal of the ESSKA, 2020: 1–9.

